# Effects of Walnut Supplementation on Short-Chain Fatty Acid Levels in Healthy Volunteers

**DOI:** 10.64898/2026.09.08.26362468

**Authors:** Thomas Petrillo, Raad Z. Gharaibeh, James J. Grady, Alexey Melnik, Alexander Aksenov, John B. Birk, Haleh Vaziri, Huijia Liu, Christian Jobin, Daniel W. Rosenberg

**Affiliations:** University of Connecticut School of Medicine, CT USA; Department of Medicine, University of Florida College of Medicine, Gainesville, FL USA; Department of Molecular Genetics and Microbiology, University of Florida, Gainesville, FL USA; Department of Chemistry, University of Connecticut, Storrs, CT USA; Division of Gastroenterology, University of Connecticut School of Medicine, CT USA; Department of Infectious Diseases and Immunology, University of Florida College of Medicine, Gainesville, FL USA; Department of Anatomy and Cell Biology, University of Florida College of Medicine, Gainesville, FL USA

**Keywords:** Dysbiosis, short-chain fatty acids, obesity, p-cresol, protein fermentation, metabolomics, clinical study, walnut, colorectal cancer, fiber

## Abstract

Gut dysbiosis poses a significant public health concern, and malnutrition plays a central role in its development. In our recent clinical trial, we explored the effects of regulated walnut intake on the levels of 11 fecal metabolites to determine whether (I) walnut consumption promotes changes in metabolite profiles consistent with reduced gut dysbiosis, and (II) whether these changes are associated with the levels of urolithin A, a bioactive metabolite produced by colonic microbes following walnut consumption. DNA was extracted from fecal samples using a DNeasy® 96 PowerSoil® Pro QIAcube® HT for 16S rRNA sequencing, and fecal short-chain fatty acids (SCFAs) and p-cresol were extracted fecal specimens by liquid chromatography/mass spectrometry (LC/MS-MS). At baseline, obese individuals (BMI > 30) had lower alpha diversity, higher p-cresol and valeric acid levels, and a higher trending *Firmicutes* to *Bacteroidetes* (F/B) ratio compared to non-obese (BMI < 30) participants, indicating a level of inherent gut dysbiosis. Walnut intake was associated with an overall 2.8% increase in alpha diversity, a 17.6% reduction in estimated marginal mean F/B ratio, and an overall reduction in three luminal proteolytic metabolites associated with gut dysbiosis: isobutyric acid, valeric acid, and isovaleric acid. Interestingly, the effects on the gut metabolome were more pronounced in obese individuals, with significant decreases observed in the same three SCFAs, as well as p-cresol. Given the known associations between these luminal proteolytic metabolites and gut dysbiosis, these results suggest that walnut intake may help to attenuate overall gut dysbiosis, particularly in obese populations.

## Introduction

Gut dysbiosis is implicated in a variety of chronic and metabolic diseases, including type 2 diabetes [1], obesity [2] and even certain cancers, including colorectal cancer (CRC) [3]. Given the persisting upward trends in these metabolic conditions [4] and an associated rise in early-onset CRC [5], there is an urgent need for accessible interventions that support a healthy gut microbiome. Diet modification has proven to be an effective strategy for preventing and managing gut dysbiosis [6], with plant-centered diets often conferring protective effects, such as enhanced diversity of beneficial gut flora [6], while diets high in red meat, saturated fat, processed foods, and refined sugar often promote inflammation and dysbiosis [6]. The gut-modulating effects of plant-based foods are largely attributed to phytochemicals, such as polyphenols, and dietary fibers, which encourage the growth of beneficial microbes while inhibiting the growth of pathogenic species [6]. Ellagic acid, a polyphenol common to nuts and berries, may also mitigate obesity-related health complications such as type 2 diabetes, non-alcoholic fatty liver disease, and atherosclerosis by regulating lipid metabolism and insulin sensitivity [7]. Likewise, insoluble fiber intake may exert similar effects by improving gut transit time and insulin sensitivity [6]. The synergistic effects of these dietary compounds are likely critical for the prevention of various chronic and metabolic diseases.

Walnuts contain an appreciable amount of fiber (1.9 g per oz [8]), and are an exceptional source of polyphenols, containing the highest concentration of all tree nuts [9]. One such polyphenol, pedunculagin, is an ellagitannin that is hydrolyzed in the stomach and converted by the gut microbiome into a series of bioactive molecules, the urolithins [10], that may may elicit protective effects in obesity [11, 12], cardiovascular disease, and cancer [7]. Previously, we showed that consuming 2-oz of whole, shelled walnuts per day increases microbial diversity and promotes a more balanced gut microbiome [13]. Given the wide array of bioactive phytochemicals present in walnuts [14], and the growth-promoting effects of prebiotic fiber on carbohydrate-fermenting microbes [15], we predicted that walnut-associated changes to the gut microbiome would be associated with beneficial changes to the gut metabolome.

Short-chain fatty acids (SCFAs), branched-chain fatty acids (BCFAs), and phenolics are bioactive metabolites produced by intestinal flora [16]. SCFAs are typically produced by saccharolytic fermentation of plant fibers and are generally beneficial to host health [17], with butyrate, acetate, and propionate serving as important regulators of gastrointestinal (GI) homeostasis and gut motility [17]. Butyrate may be especially beneficial within the context of CRC pathogenesis, given its association with enhanced gut barrier function and its known ability to suppress tumor growth and NF-κB activity in HT-29 cancer cells [17]. Conversely, BCFAs, ammonia, and phenolics are produced by proteolytic fermentation of branched-chain amino acids and peptides [16, 18], and may elicit detrimental effects on host health, potentially increasing the risk of obesity, type 2 diabetes, and CRC [16]. In the current study, the fecal levels of eleven luminal metabolites, including BCFAs and phenolics (proteolytic metabolites), were measured in the feces of a subgroup of 21 healthy participants to determine whether the reported beneficial effects of walnut intake [10, 13, 19] may extend to the gut metabolome.

## Materials and Methods

### Study design

The study design for this study has been reported earlier [10]. Participants were screened at visit 1 to ensure eligibility [10]. After patient selection, 47 subjects were enrolled into the study. A total of 39 healthy patients, 21 females and 18 males, 50 - 65 years of age, completed the study. Consenting participants underwent a one-week run-in where they abstained from consuming dietary supplements and ellagitannin-containing foods such as nuts, pomegranates and berries [10]. Afterwards, participants consumed 56 grams (2-oz) of whole, shelled walnuts per day for 3 weeks. Urine and fecal samples were collected at the end of the one-week run-in to serve as a baseline, and again at the end of the 3-week dietary intervention. Nutrient compliance was assessed *via* 3-day dietary records completed during the week of sample collection. Block Food Frequency Questionnaires (BFFQs) were also included to estimate participant dietary behavior during the past 12 months [10]. As outlined earlier [10], the informed consents, collection of clinical data, demographic data, and study protocols followed strict institutional guidelines. Study compliance was also assessed by measuring the levels of urinary creatinine-normalized urolithin A (ng/mg) immediately following the one-week washout period. Urolithin levels at this time-point should be close to or at zero, assuming all ellagitannin-containing foods were properly avoided. BMI levels were calculated for all participants based on weight and height measurements acquired during the initial office visit. By convention, participants with BMI values ≥ 30 were considered obese, while those with BMI values < 30 were considered non-obese. (*Clinicaltrials*.*gov*, *number*: *<u>NCT04066816</u>)*.

The methods used to extract fecal DNA and perform 16S RNA sequencing have been reported previously [13]. Briefly, microbial DNA from stool samples from all 39 participants were extracted using the DNeasy® 96 PowerSoil® Pro QIAcube® HT (QIAGEN). PCR products were analyzed on a 2% agarose DNA gel, and qPCR products were quantified using a KAPA Library Quantification Kit (KAPA Biosystems). For 16S RNA gene sequencing, the DADA2 pipeline was used to process de-multiplexed reads. Barcoded primer pairs 341F (5’ = - CCTACGGGNGGCWGCAG-3’) and 785R (5’ = -GACTACHVGGGTATCTAATCC-3’ =) were used to amplify the 585 base-pair of 16S RNA V3-V4 hypervariable region.

To establish our sample cohort for fecal metabolomics analyses, the 39 patients who completed the study were separated into subgroups based upon their capacity to form urolithin A, measured in the urine. For the subgroup analysis, unbiased, simple random samples were used to randomly select 10 participants with high urolithin A-producer status and 11 participants with low urolithin A-producer status, with means of 21,138 (range: 7,553 to 72,740) ng/mg and 182 (range: 0 to 435) ng/mg creatinine in the high and low groups, respectively.

### Metabolomic analyses of fecal and urine samples

After collection of fecal aliquots, samples were stored at -80°C until further analysis. LC/MS-MS was used to quantify fecal urolithin A levels as described earlier [10]. Briefly, a Vanquish UPLC (Thermo Fisher Scientific) was used to inject samples onto an Orbitrap Exploris 480 (Thermo Fisher Scientific) mass spectrometer (MS) in negative polarity, equipped with HESI-II probe sources, as described in our recent publication [20].

Urine samples were collected from all 39 participants at the end of the one-week wash-out and immediately prior to the colonoscopy procedure. The first urine collection enabled sufficient depletion of systemic urolithin metabolites prior to walnut supplementation, while providing further validation of patient compliance. Spot urine samples collected by patients were aliquoted and stored at -80°C within 24 hours of collection. To accurately quantify urinary urolithin levels, samples were subjected to overnight hydrolysis with arylsulfatase and beta-glucuronidase to release free urolithins that would otherwise be bound as glucuronides (∼95%), as well as sulfates, sulfoglucuronides, and diglucuronides [21, 22].

For GC-MS data acquisition and processing of SCFAs (listed under **Supplemental Table 1**), 1-μL aliquots of fecal supernatants were directly injected onto an Agilent 6890 GC interfaced to a mass spectrometer LECO BT for electron ionization GC-MS. The GC utilizes a 30m ZB-FFAP column (0.25 mm i.d., 0.25 μm film thickness) for metabolite separation with 1.2mL/min constant He flow. The oven temperature program initiates at 50°C rising to 240°C at 10°C/min. No noticeable carryover was observed over the entire injection sequence for all the studies. Also, no increased contamination that necessitates liner change was observed, indicating that possible lint particulate traces from wipe material did not contribute to observable analytical interference. The data were then deconvoluted with the MSHub algorithm [23]. The experimental spectra were searched against the NIST 2023 library with ≥80% spectral match defining putative identifications, with the retention times falling within <0.01 min of the corresponding reference standards. Targeted analysis of SCFAs and p-cresol was performed by using Agilent Masshunter software.

### Statistical analysis

The software open software package, Jamovi, was used to perform all statistical analyses. Statistical models for analysis of the metabolomics data were chosen according to data skewness, normality, and homogeneity. Distribution descriptives were used to identify data skew. Log₂-fold changes (log₂FC) were calculated using the formula Log₂(post/pre). Metabolomics data were log_10_-transformed if skewness was greater than or equal to 1.0. Assumption checks were used to determine the most appropriate paired-sample tests. Shapiro-Wilk tests were used to determine whether data was normally distributed. Shapiro-Wilk p = <0.05 indicated a non-normal distribution, necessitating the use of Mann-Whitney U or Wilcoxon W-test rather than Student’s t test. Linear mixed models were used to test for differential effects between subgroups, such as BMI, or to adjust for factors such as daily fiber intake and daily protein intake. Fixed terms included timepoint, BMI, and timepoint x BMI, while intercept – subject ID was used as a random term. For paired-sample tests, only the 38 participants with data from both timepoints were included. Conversely, all 39 participants were included in linear mixed modeling analyses. To compare abundancies of microbial genera across timepoints, the equation log_10_(value + 0.5) was used to ensure that every datapoint would be included in the analysis even if raw abundance equaled zero. To calculate percent changes in the abundance of microbial genera, the equation ((post – pre)/pre) * 100 was used. For our aggregated analysis of 11 saccharolytic genera, the abundances of all 11 taxa were summed for each participant. All post-hoc analyses were adjusted using Tukey-HSD.

## Results

### Fecal metabolite levels and associated changes to gut microbiome

We compared the levels of 11 metabolites in fecal samples obtained from a subgroup of 21 patients, before and after a 3-week walnut diet supplementation. The bar plots represented in **Figure 1A-D** show the distribution levels (log_10_-transformed) of four of these metabolites (a full list of analytes is shown in **Table 1**, where a negative log_2_-fold change indicates that levels decreased post-walnut, while a positive log_2_-fold indicates that levels increased post-walnut.). Since the luminal formation of these metabolites may be affected by protein and fiber intake, we adjusted for these covariates in our linear mixed models using data from participant BFFQs collected at study entry. The levels of the SCFAs isobutyric acid (log₂FC = -0.71, p = 0.005), valeric acid (log₂FC = -1.05, p = 0.031), and isovaleric acid (log₂FC = -0.66, p = 0.014), were significantly reduced after walnut ingestion (**Table 1**). p-Cresol levels also trended downward post-walnut but were not significantly reduced (log₂FC = -0.79, p = 0.072).

**Figure 1.**
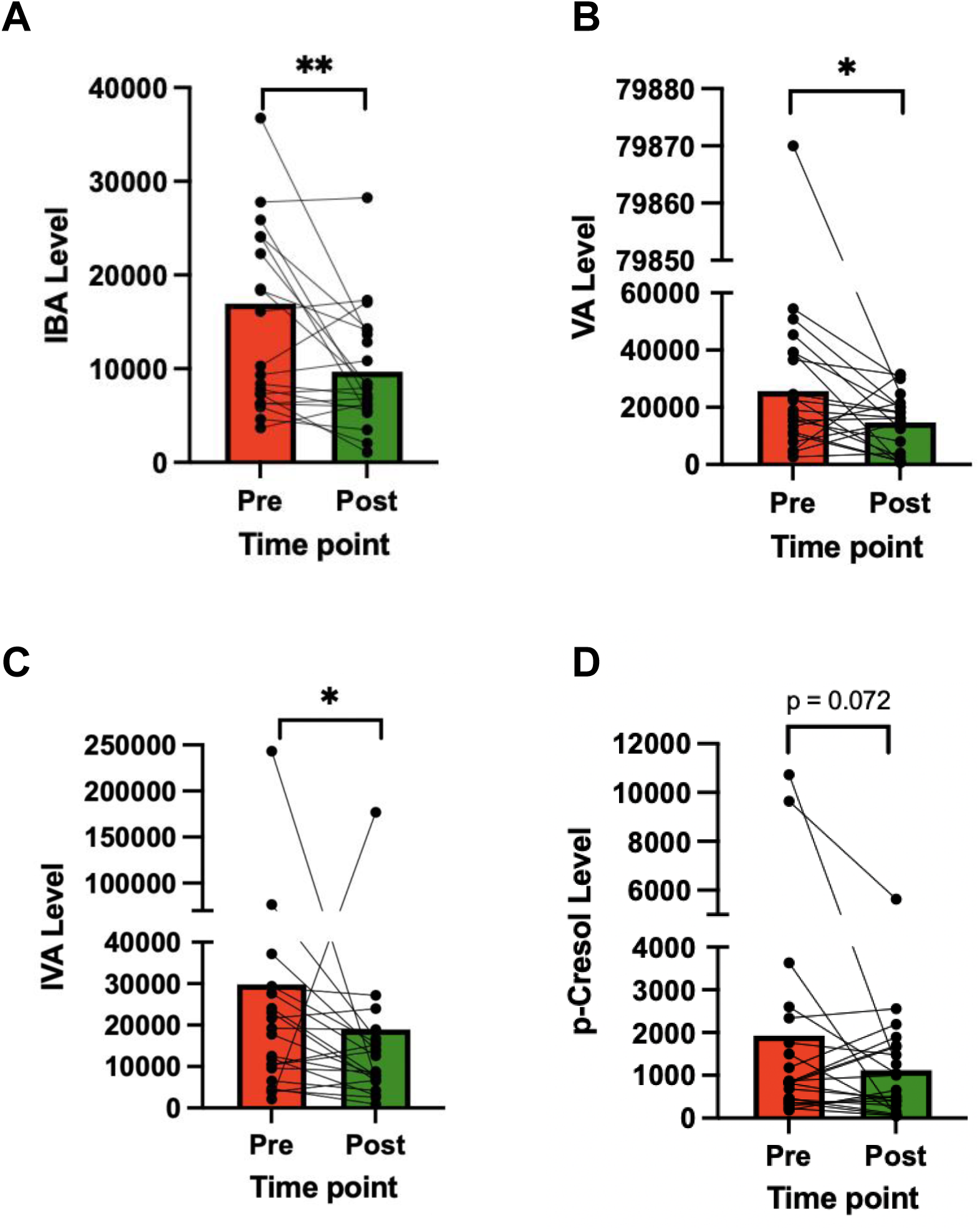
The effect of walnut supplementation on fecal metabolite levels in a subgroup analysis of 21 study participants. Comparison of fecal levels of four metabolites, A) isobutyric acid, B) valeric acid, C) isovaleric acid and D) p-cresol, before and after walnut supplementation. Each data point represents an individual patient (paired values, pre- and post-walnut). Paired samples Student’s t-tests were performed using log_10_-transformed data to adjust for numerical skew. A p-value less than 0.05 was considered statistically significant (*), while a p-value less than 0.01 was considered highly significant (**). Raw values were plotted for ease of visualization, while log-transformed values were used for statistical analysis. Walnut intake significantly reduced the levels of 3/4 fecal metabolites: isobutyric acid (p = 0.005), valeric acid (p = 0.031), and isovaleric acid (p = 0.014).

**Table 1.**
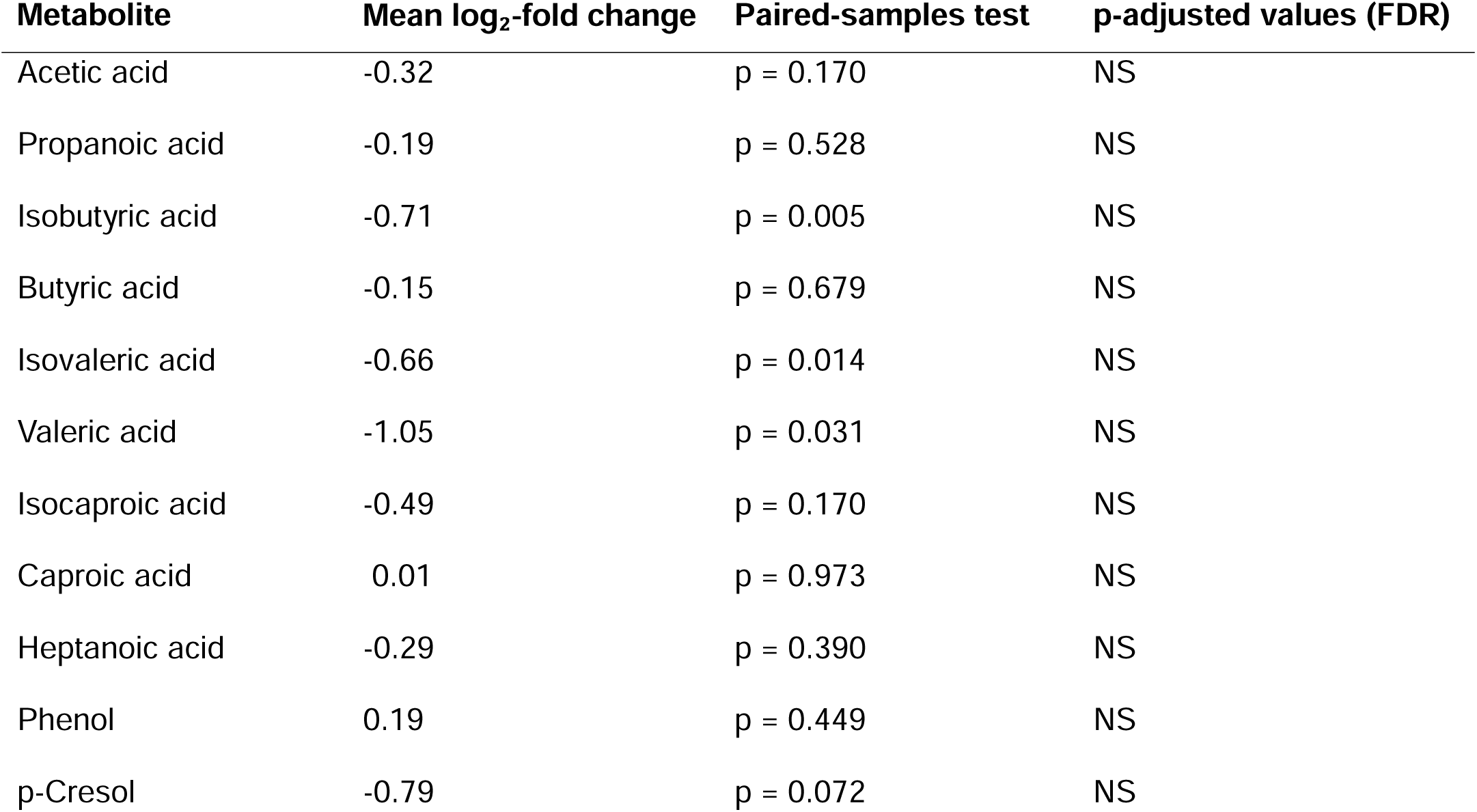
Log_2_-fold change and statistical significance for changes in the fecal levels of 11 metabolites during a three-week walnut supplementation.

| Metabolite | Mean log<sub>2</sub>-fold change | Paired-samples test | p-adjusted values (FDR) |
| --- | --- | --- | --- |
| Acetic acid | -0.32 | p = 0.170 | NS |
| Propanoic acid | -0.19 | p = 0.528 | NS |
| Isobutyric acid | -0.71 | p = 0.005 | NS |
| Butyric acid | -0.15 | p = 0.679 | NS |
| Isovaleric acid | -0.66 | p = 0.014 | NS |
| Valeric acid | -1.05 | p = 0.031 | NS |
| Isocaproic acid | -0.49 | p = 0.170 | NS |
| Caproic acid | 0.01 | p = 0.973 | NS |
| Heptanoic acid | -0.29 | p = 0.390 | NS |
| Phenol | 0.19 | p = 0.449 | NS |
| p-Cresol | -0.79 | p = 0.072 | NS |

Using 16S rRNA sequencing data collected from our previous study [13], we generated a summary table of proteolytic, saccharolytic, and mucin-fermenting genera that shifted in abundance following walnut intake in the entire study population. **Table 2** highlights the observation that walnut intake was associated with significantly increased abundance of multiple saccharolytic genera and decreased abundance of proteolytic and mucin-fermenting genera [13]. These data provide important mechanistic context to the walnut-associated changes we observed in fecal metabolite profiles.

**Table 2.** Metabolic phenotypes of microbes altered by 21 days of walnut supplementation.

| Genera/Species | Metabolism | Effect of Walnut |
| --- | --- | --- |
| *Anaerococcus* | Proteolytic | Decreased |
| *Clostridium innocuum* | Mixed; saccharolytic and proteolytic | Decreased |
| *Romboutsia* | Mixed; saccharolytic and proteolytic | Decreased |
| *Rumminococcus gnavus* | Mucin-fermenting | Decreased |
| *Roseburia* | Saccharolytic | Increased |
| *Eubacterium eligens* | Saccharolytic | Increased |
| *Lachnospira* | Saccharolytic | Increased |
| *Butyricicoccus* | Saccharolytic | Increased |
| *Gordonibacter* | Polyphenol-fermenting (UroA producer) | Increased |

### The effect of obesity status on fecal metabolite levels

We next stratified the data based on the recorded BMI of each study participant and reevaluated the levels of all 11 fecal metabolites, before and after walnut supplementation. These data were adjusted for total daily fiber and protein intake from the BFFQ. Interestingly, significant walnut-related decreases were observed for the same four metabolites (isobutyric acid, valeric acid, isovaleric acid, and p-cresol) described above (**Fig. 1A-D**), but this effect was only present in obese individuals (**Fig. 2A-D**). The results for all 11 metabolites are shown in **Supplemental Table 2**. Additional statistical comparisons were performed to determine whether high and low-BMI groups had different levels of these metabolites at baseline (**Fig. 3A-D**). The fecal levels of valeric acid (B) and p-cresol (D) were significantly elevated in obese individuals pre-walnut (p = 0.036 and 0.022, respectively), while no significant differences were observed in the levels of isobutyric acid (A) or isovaleric acid (C).

**Figure 2.**
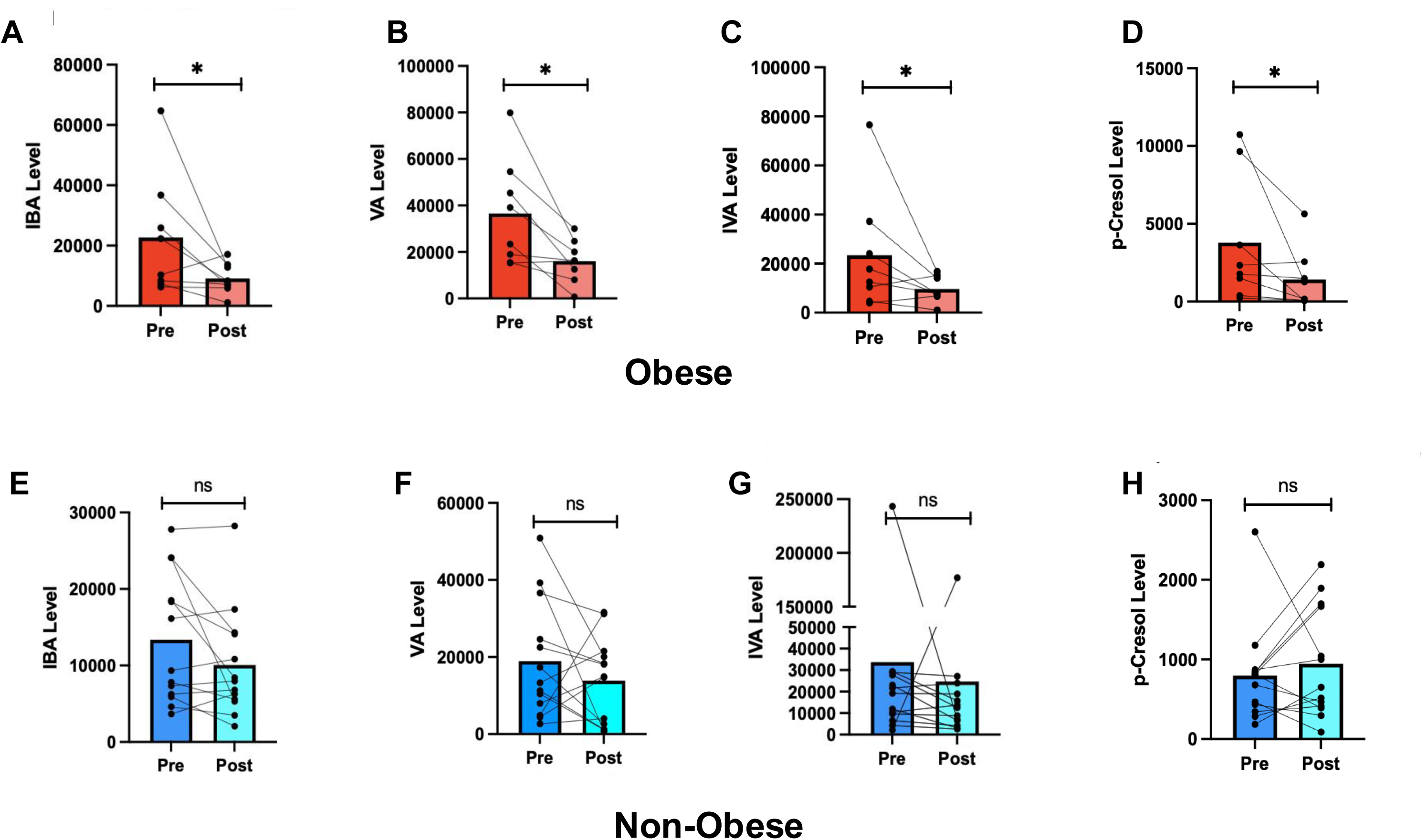
The effects of walnut supplementation on fecal metabolite levels in a subgroup analysis of obese and non-obese study participants. The fecal levels of four metabolites, (A,E) isobutyric acid, (B,F) valeric acid, (C,G) isovaleric acid and (D,H) p-cresol, before and after walnut supplementation. Paired samples Student’s t-tests were performed using log_10_ transformed data to adjust for numerical skew. A p-value less than 0.05 was considered statistically significant (*). In obese individuals (A-D), walnut intake significantly reduced the levels of all four fecal metabolites: isobutyric acid (p = 0.028), valeric acid (p = 0.016), isovaleric acid (p = 0.043), and p-cresol (p = 0.015). No significant changes were observed in non-obese individuals. (E-H). Sample sizes were n = 8 and n = 13 for obese and non-obese groups, respectively.

**Figure 3.**
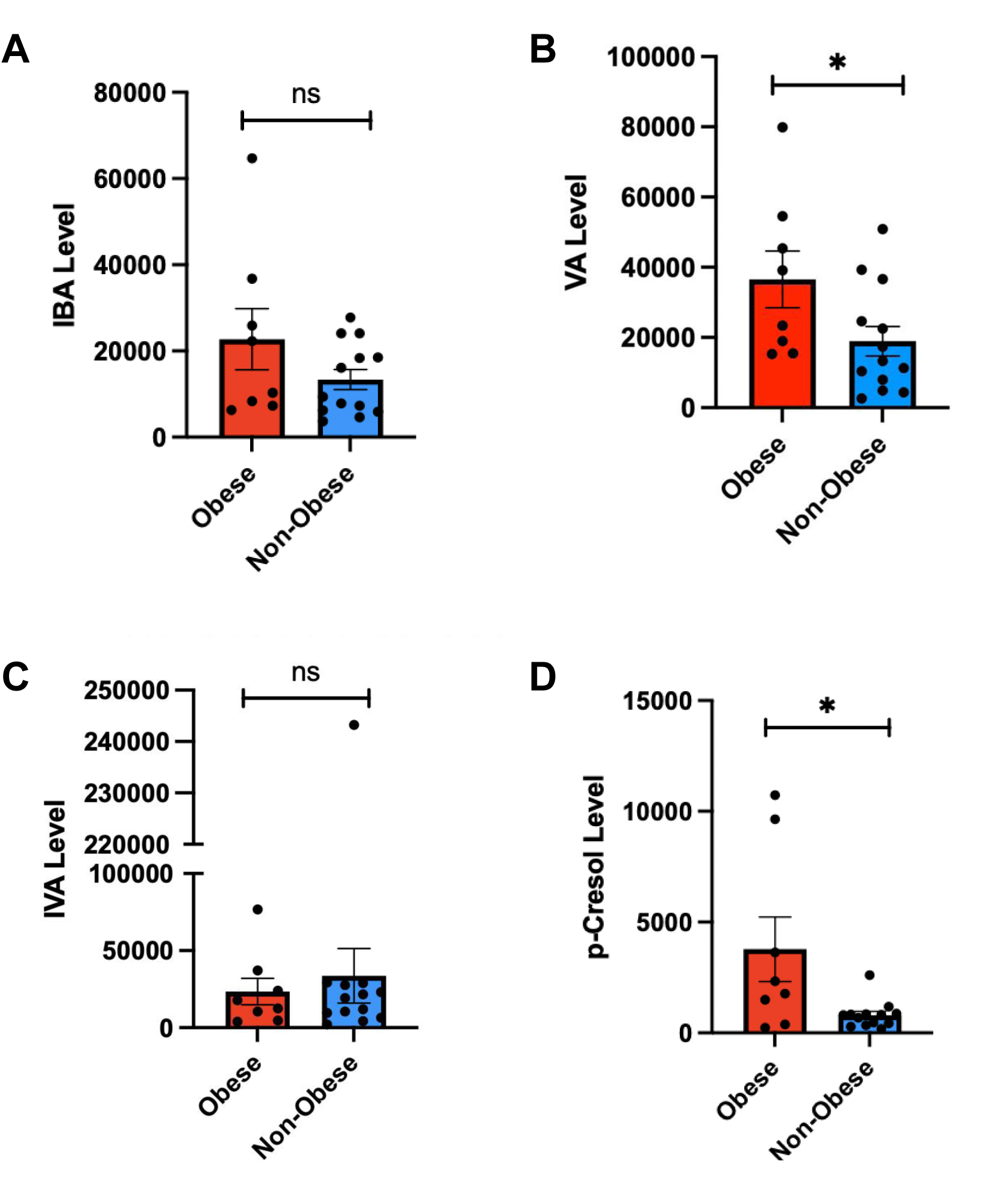
Baseline fecal metabolite levels in a subgroup analysis of obese and non-obese study participants prior to walnut supplementation. The fecal levels of four metabolites, A) isobutyric acid, B) valeric acid, C) isovaleric acid and D) p-cresol, before and after walnut supplementation. Independent samples tests were performed using log_10_-transformed data to adjust for numerical skew. A p-value less than 0.05 was considered statistically significant (*). Valeric acid and p-cresol levels were significantly higher (p = 0.036 and p = 0.022) in the obese group compared to the non-obese group. Sample sizes were n = 8 and n = 13 for obese and non-obese groups, respectively.

To further explore whether high- and low-BMI individuals responded differently to walnut, we next compared the responses of obese and non-obese individuals to walnut intake using linear mixed modeling analyses. Data was log-transformed to adjust for skewness. As shown in **Figure 4A-E**, a significant reduction in isobutyric acid levels (A) was limited to obese individuals, post-walnut (p = 0.014). However, the combined effect of timepoint and BMI did not reach significance (p = 0.102), possibly due to the relatively small sample sizes of these groups. Interestingly, post-walnut isobutyric acid levels were normalized across both BMI groups, suggesting that walnut supplementation may have mitigated the effects of obesity on isobutyric acid levels. Similar effects were observed for p-cresol (D), with a significant overall effect of timepoint in obese individuals (p = 0.006), no significant change in non-obese individuals, and a shift toward relatively comparable p-cresol levels between high- and low-BMI groups post-walnut. However, unlike the results shown for isobutyric acid, the overall effect of walnut supplementation on p-cresol levels was significantly different between obese and non-obese groups (p = 0.003), with the higher BMI group showing a 62.8% reduction post-walnut compared to the non-obese group, which trended upward post-walnut. Similar, although non-statistically significant trends, were observed for the post-walnut levels of the SCFAs, valeric acid (B)and isocaproic acid (E). Mean valeric acid levels trended downward in both BMI groups post-walnut, with relative reductions of 56.1% and 26.8% in obese and non-obese groups, respectively.

**Figure 4.**
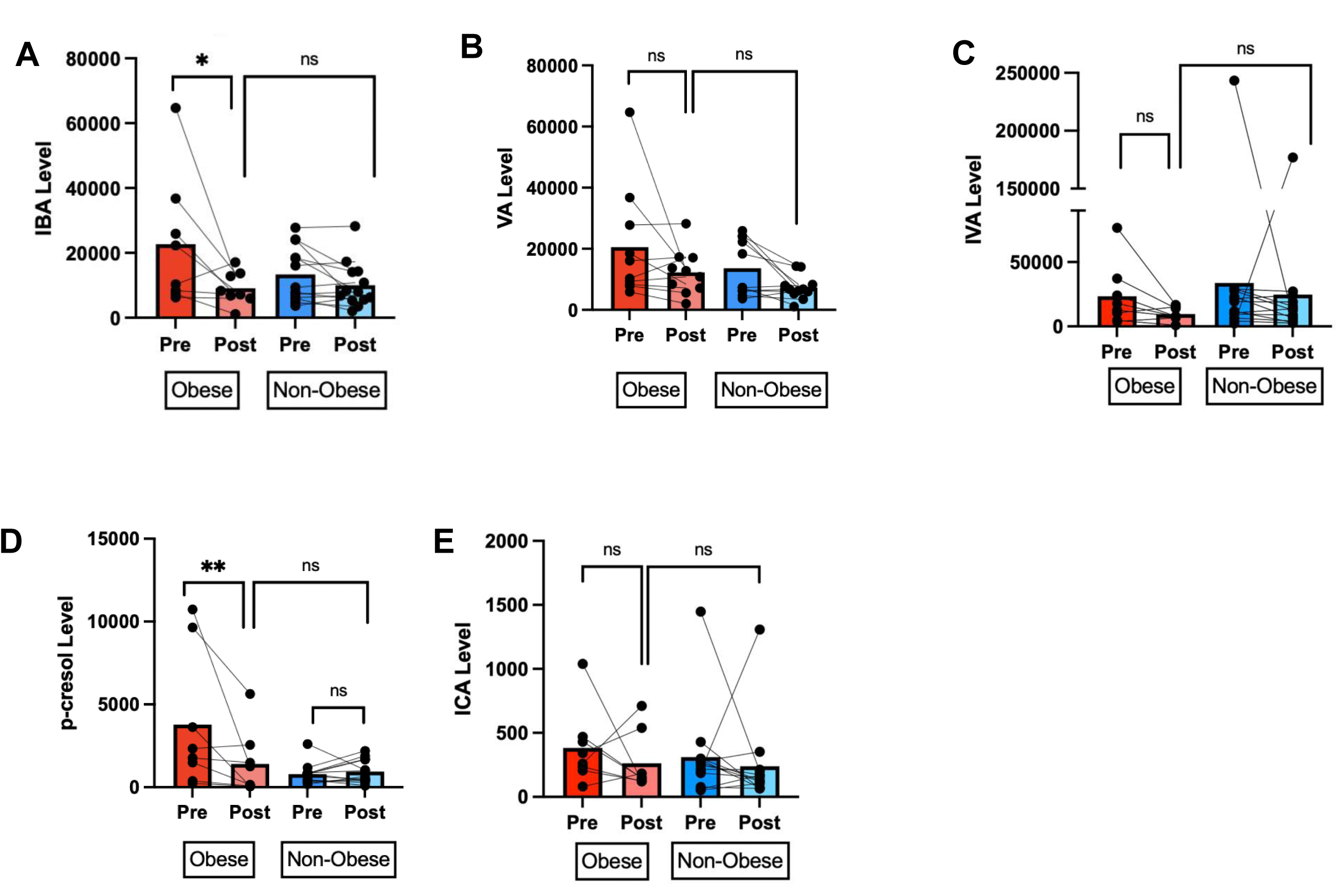
Comparison of changes in fecal metabolite levels in a subgroup of obese *vs*. non-obese study participants. To determine whether low and high BMI groups respond differently to walnut ingestion, the fecal levels of five metabolites, (A) isobutyric acid, B) valeric acid, C) isovaleric acid, D) p-cresol and (E) isocaproic acid, before and after walnut supplementation were compared. Linear mixed models were used to assess the combined effects of walnut intake and BMI. Data were log_10_-transformed to adjust for numerical skew. A p-value less than 0.05 was considered statistically significant (*). Sample sizes were n = 8 and n = 13 for obese and non-obese groups, respectively.

### The Effect of Obesity Status on Markers of Dysbiosis

To understand why obese and non-obese individuals may have responded differently to walnut intake, we explored the effect of obesity status on a panel of markers of dysbiosis. Changes in alpha diversity and F/B ratio in obese and non-obese subgroups from the full 39-participant study pool were analyzed using linear mixed models. **Figure 5** compares the alpha diversity (A) and F/B ratios (B) calculated using RNA-sequencing data from fecal samples obtained during the pre- and post-walnut intake periods. The overall effect of obesity status on alpha diversity was significant (p = 0.006), with obese participants having lower alpha diversity compared to non-obese participants at baseline (p = 0.044), while no significant difference in alpha diversity was detected between these BMI groups post-walnut (p = 0.056). The overall effect of timepoint was significant (p = 0.028), with walnut intake associated with a 2.8% increase in overall alpha diversity among all participants. Subgroup *post-hoc* analyses of obese and non-obese individuals revealed no significant changes in alpha diversity (p = 0.470 and 0.221, respectively).

**Figure 5.**
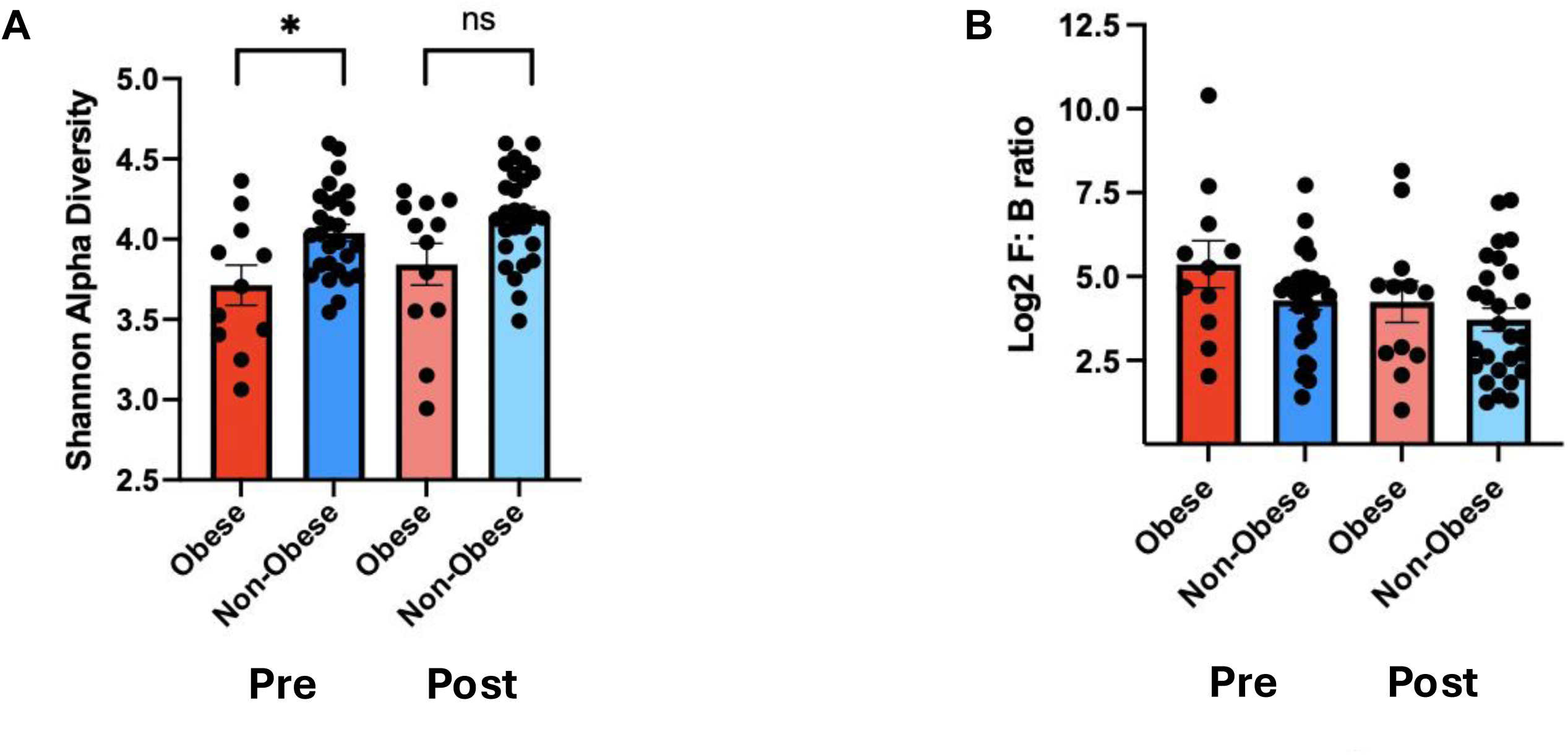
The effect of walnut intake on alpha diversity and F/B ratio in a subgroup analysis of obese and non-obese study participants. (A) Alpha diversity and (B) F/B ratios were calculated using 16S rRNA-sequencing data from fecal samples obtained during the pre- and post-walnut intake intervals, as described under Materials and Methods. Linear mixed models with the Tukey *post-hoc* test were used to determine the effect of walnut intake and obesity status on alpha diversity and log_2_-transformed F/B ratios. A p-value less than 0.05 was considered statistically significant (*). Sample sizes were n = 12 and n = 27 for obese and non-obese groups, respectively.

For F/B ratio, a linear mixed model showed a significant overall effect of timepoint (p = 0.030), with a 17.6% reduction in estimated marginal mean F/B ratio post-walnut. The F/B median was reduced from 4.60 to 3.58. While the overall effect of BMI was not significant (p = 0.128), the obese group had higher trending F/B ratios at both timepoints compared to the non-obese group. The combined effect of timepoint and BMI was also not significant (p = 0.473), suggesting that changes in F/B ratio were not significantly different between high and low BMI groups.

### The effect of obesity status on microbial community structure

In the following analysis, we compared the effect of participant BMI on walnut-associated changes to the fecal microbiome profiles reported in our recent study [13], using data from all 39 participants. While not statistically significant, *Peptostreptococcus* levels trended downward in both BMI groups post-walnut (**Supplemental Fig. 2A**), although the obese individuals showed a greater reduction (74.2 *vs*. to 58.6%, respectively; **Supplemental Table 3**). The relative abundance of *CSS1* also trended downward in the obese group, post-walnut (**Supplemental Figure 2B**), with a relative reduction of 52.8% (obese n = 11) (**Supplemental Table 3**). Interestingly, *CSS1* levels trended upward in the non-obese group, post-walnut, with a relative increase of 27% (n = 27). The mean abundance of 11 saccharolytic genera altered by walnut intake [13] also trended upward in both obese and non-obese BMI groups post-walnut, with relative increases of 47.1% (p = 0.304) and 146.4% (p = <0.001), respectively. Moreover, the relative abundances of three saccharolytic genera, *Lachnospiraceae UCG001*, *Butyricicoccus*, and *Eubacterium eligens*, trended higher in non-obese individuals compared to obese individuals overall (**Supplemental Table 4**).

### The effect of Urolithin A-producer status on fecal metabolite levels

The capacity of individual subjects to form urolithin A was also associated with significant changes in the levels of several fecal metabolites post-walnut. These results are shown in **Figure 6A-E**, and the corresponding data can be found in **Supplemental Table 1**. As with our prior analyses, we used linear mixed-models with log-transformation to adjust for skewed data. Given the effect of obesity status on the levels of these metabolites, participant BMI (categorical) was included as a covariate to adjust for confounding. Significant decreases in the levels of p-cresol (A), isobutyric acid (C), and valeric acid (D) were observed in the low urolithin A producers, post-walnut (p = 0.001, p = 0.032, and p = 0.037, respectively), while no significant changes were observed in the high urolithin A producers. Concurrently, the effect of urolithin A-producer status was significant for phenol (A) and p-cresol (B) (p = 0.031 and p = 0.002), indicating that differences in urolithin A formation were associated with SCFA metabolism. Furthermore, the combined effect of timepoint and urolithin A-producer status was not significant for phenol (p = 0.108) but was significant for p-cresol (p = 0.011), the latter indicating that changes in fecal p-cresol levels were significantly different between high *versus* low urolithin A producers. Despite the apparent lack of urolithin A-associated changes in fecal phenol levels, *post-hoc* analysis revealed that post-walnut phenol levels were significantly lower in the high urolithin A producers compared to low urolithin A producers (p = 0.033). A separate examination of the effect of urolithin A-producer status on p-cresol levels in obese and non-obese groups can be found in **Supplemental Figure 1A, B**.

**Figure 6.**
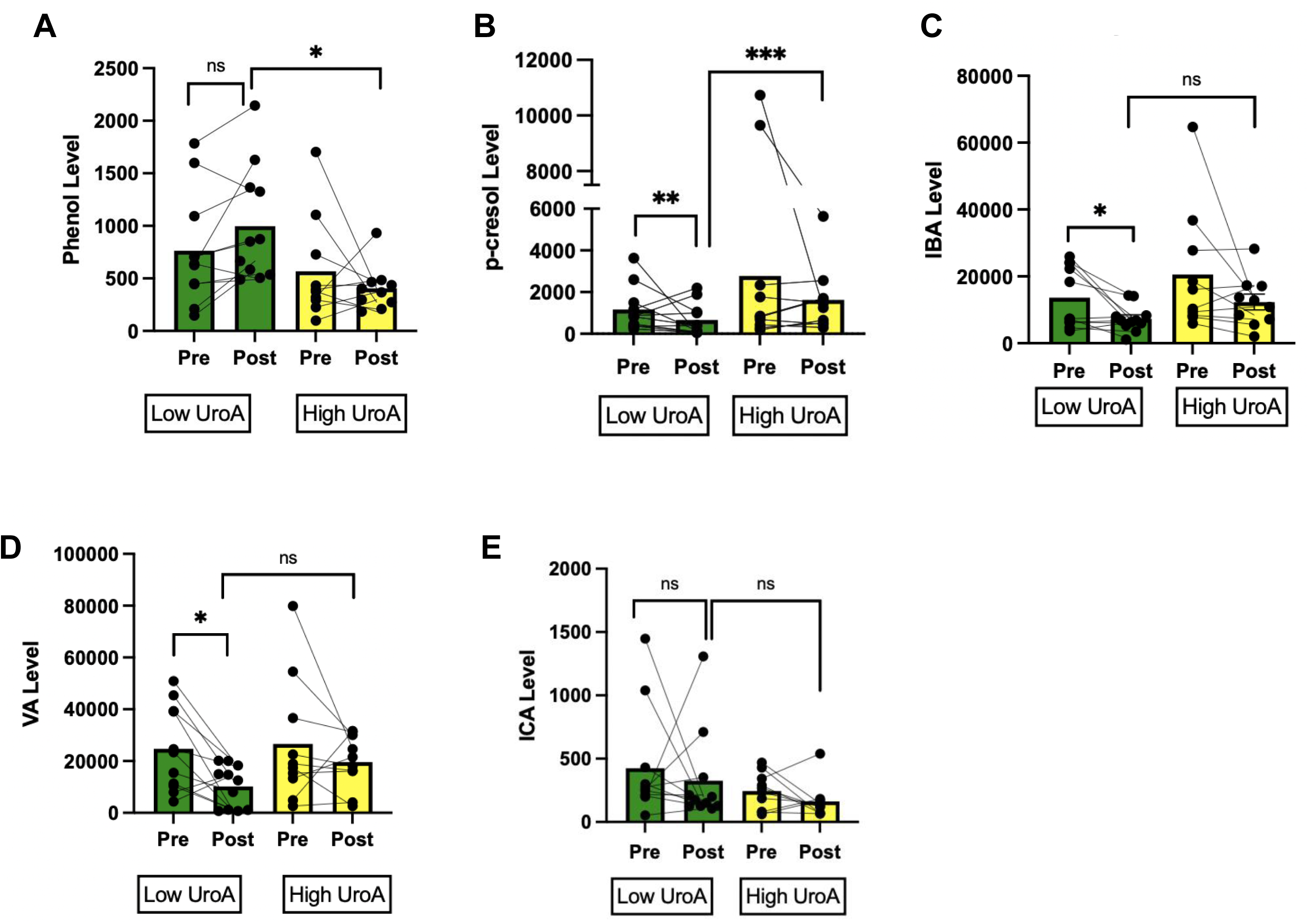
The effect of urolithin A-producer status (low *vs*. high) on walnut-related changes to fecal metabolite levels in a subgroup analysis of study participants. The fecal levels of five metabolites, (A) phenol, (B) p-cresol, (C) isobutyric acid, (D) valeric acid and (E) isocaproic acid, were measured before and after walnut supplementation to determine whether urolithin A levels in the urine correlates with changes in fecal metabolite levels. Linear mixed models were used to assess the combined effect of walnut intake and urolithin A-producer status (high *vs.* low). Data were log_10_-transformed to adjust for numerical skew. Participant BMI was included as a covariate to adjust for potential confounding. A p-value less than 0.05 was considered statistically significant (*). Sample sizes were n = 12 and n = 10 for low- and high-urolithin A groups, respectively.

## Discussion

Branched chain fatty acids (BCFAs), like isobutyric and isovaleric acid, and other luminal metabolites, like p-cresol, phenol, ammonia, and valeric acid, are products of intestinal putrefaction [16]. Elevated fecal levels of these products may indicate excessive proteolysis and gut dysbiosis [16, 24–27], making them potentially valuable biomarkers for GI-related diseases. Among these metabolites, phenol, ammonia, and p-cresol have been shown to directly impair intestinal barrier function by increasing tight-junction permeability [28, 29]. In fact, p-cresol may be especially important given its reported genotoxic effects on colonic epithelial cells [24, 29] and its role as a uremic toxin implicated in the development of renal and cardiovascular complications in patients with chronic kidney disease [29]. While the direct effects of BCFAs and valeric acid on the colonic mucosa are incompletely understood, elevated fecal/urinary levels of these metabolites, and p-cresol, have been associated with overall increases in obesity and CRC risk [30–32]. Given the association between obesity and gut dysbiosis [4], diet interventions that reduce colonic putrefaction may disproportionately benefit obese individuals and those suffering from obesity-related metabolic disorders.

Walnuts may be particularly well-suited for reducing colonic putrefaction in obese individuals, potentially reducing their elevated risk of gut dysbiosis and CRC. The association between putrefaction end products and obesity [32] may be explained in part by altered gut transit time and colonic motility observed in obesity [33]. This dysfunction may impair nutrient digestion and absorption [34], potentially allowing more undigested protein to reach the colon where it may be fermented [33]. In fact, slower colonic transit time is associated with GI carbohydrate depletion and subsequent increases in putrefaction, leading to greater production of BCFAs and ammonia [33]. Walnut-based insoluble fibers may directly counter these effects by improving gut transit time [6] and increasing carbohydrate fermentation [35], supporting a more balanced gut microbiome, and by extension, a more balanced gut metabolome.

As shown in **Figure 1**, three-week supplementation with walnut significantly reduced the fecal levels of isobutyric acid, valeric acid, and isovaleric acid in a sub-sampling of 21 participants enrolled in this pilot clinical trial. Further stratification by BMI revealed a greater reduction in all three metabolites, as well as p-cresol, the latter effect of which was limited to obese participants (**Fig. 2).** These results were adjusted for total daily protein and fiber intake data obtained from BFFQs, suggesting that differences in the intake of these nutrients between BMI groups did not bias these results. While no significant effects were observed in the non-obese group (**Fig. 2**), the average levels of isobutyric acid, valeric acid, and isovaleric acid trended downward post-walnut, implying potentially modest changes in fecal metabolite profiles.

At baseline, obese participants had significantly higher levels of valeric acid (**Fig. 3**) and p-cresol (**Fig. 3**) compared to their non-obese counterparts. These findings support an earlier study by *Tihonen et al.*, which reported increased levels of phenolic compounds and valeric acid in the feces of obese volunteers compared to healthy-weight controls [32]. However, unlike this earlier study [32], we found no significant differences in baseline BCFA levels between obese and non-obese groups, potentially reflecting the variability of host microbiome composition among obese individuals. Importantly, post-walnut fecal levels of BCFAs, p-cresol, and valeric acid were similar between obese and non-obese groups (**Fig. 4**), suggesting that for obese participants, walnut intake may have normalized the production of these metabolites towards a concentration range that is consistent with healthy weight status. These results may be of significance given the proposed link between putrefaction products and metabolic diseases, including obesity, type 2 diabetes, and non-alcoholic fatty liver disease [16, 36–38].

The observed walnut-associated changes to fecal metabolite profiles are most likely dependent upon diet-related changes to the host microbiome. To examine associations between microbial composition and SCFA profiles, 16S rRNA sequencing data from all 39 participants obtained from our recent study [13] was re-analyzed following BMI stratification (12 obese and 27 non-obese). At baseline, obese individuals had significantly lower alpha diversity and a higher trending F/B ratio compared to their non-obese counterparts, indicating a level of inherent gut dysbiosis (**Fig. 2**). Walnut intake improved overall alpha diversity and F/B ratios (**Fig. 2**), and significantly reduced the levels of 15 genera, including three that contain proteolytic species: *Anaerococcus* [39, 40], *Romboutsia* [41–43], and *Clostridium [innocuum]* [44]. Members of the genus *Anaerococcus* are broadly proteolytic [40], with species like *Anaerococcus prevotii* producing ammonia from threonine and serine [39]. Similarly, all clades of *clostridium innocuum* possess multiple complete amino acid-metabolizing pathways [44]. The genus *Romboutsia* is metabolically diverse, containing several species that utilize carbohydrates and others that ferment amino acids [41]. Accordingly, it is possible that the decreases we observed in the levels of fecal proteolytic metabolites are due, in part, to changes in the abundance of these species/genera.

Walnut-associated reductions in proteolytic genera may be beneficial to the host microbiome. Although the effects of *Romboutsia* on colonic health are not well understood, a large-scale quantitative profiling study of various bacterial strains found that elevated fecal levels of *Anaerococcus vaginalis* and *Peptostreptococcus anaerobius,* a proteolytic species [45] within the same taxonomic family as *Romboutsia* [46], were associated with intestinal inflammation and CRC risk [47]. Similarly, *C. innocuum* has been implicated in the exacerbation of *C. difficile*-associated inflammatory bowel disease (IBD) [48], a known risk factor for CRC [49]. Future studies examining whether *Anaeroccocus*, *Romboutsia,* and *C. innocuum* produce BCFAs and phenolics in carbohydrate-deficient culture could provide important mechanistic insight into their potential pathophysiology. The relative abundance of another potentially detrimental species, *Ruminococcus* [*gnavus*], was also significantly reduced after walnut [13]. *R. gnavus* is a mucin-fermenting bacterium often associated with Crohn’s Disease [50]. Given that low-fiber intake is associated with increased fermentation of mucins [51], it is likely that increased fiber intake from walnut contributed to the reduction in the abundance of this species. Collectively, the reduction in these taxa following walnut intake may indicate improved overall gut health.

Following participant stratification by BMI, our results further suggested that walnut consumption may have reduced the abundance of luminal proteolytic genera. These data are shown in **Supplemental Table 2.** Walnut intake was associated with a non-significant reduction in the levels of *peptostreptococcus* in both non-obese and obese subjects, with the high-BMI group having the greatest reduction post-walnut. Furthermore, walnut intake appeared to minimize differences in the levels of *peptostreptococcus* between BMI groups, mirroring the effects of walnut intake on the levels of several proteolytic metabolites discussed earlier. The abundance of another proteolytic genera, *clostridium senso stricto 1,* also trended downward post-walnut, although this result was also not significant (p = 0.814). Meanwhile, the overall abundance of 11 saccharolytic microbes reported in our recent study [13] was significantly increased in non-obese participants after walnut, and trended upwards in obese participants after walnut, indicating that walnut consumption improved overall host microbiome composition. Non-significant statistical trends also indicated that obese participants had higher baseline levels of *peptostreptococcus* compared to non-obese subjects. This observation is consistent with earlier studies showing that *peptostreptococcus* levels are elevated in the gut of obese individuals with type 2 diabetes [52]. *Peptostreptococcus anaerobius* has been shown to be associated with increased risk of CRC, although its cancer-promoting mechanisms are poorly understood [53].

In addition to reducing the abundance of potentially detrimental proteolytic microbes, walnut intake appears to increase the abundance of beneficial saccharolytic microbes. In our recent study, walnut intake was associated with increased fecal levels of multiple saccharolytic and polyphenol-fermenting genera [13], several of which are associated with increased production of butyric acid, a sign of improved gut health [54]. These results support an earlier study by *Holscher et. al* demonstrating that walnut consumption increased the relative abundance of butyrate-producing taxa [55]. Interestingly, results from the current study further suggest a potential association between the expansion of *Gordonibacter* [13], a known producer of urolithins [56], and changes in gut proteolysis. After walnut, the high urolithin A producers had significantly lower fecal phenol levels compared to the low producers (**Fig. 6**), while no significant differences between these groups were observed at baseline. A similar, albeit non-significant trend, was observed for the BCFA, isocaproic acid (**Fig. 6**). Furthermore, the levels of two beneficial SCFAs, butyric and caproic acid, trended higher in high urolithin A producers compared to the low producers (**Table 1**), although these results were not statistically significant. Consistent with our previous study [10], these data suggest that higher urolithin A levels may be beneficial to colon health. Paradoxically, the levels of three proteolytic metabolites, p-cresol, isobutyric acid, and valeric acid, were all significantly reduced in the low urolithin A-producer group post-walnut, an effect that was not observed in the high urolithin A producers (**Fig. 6**). Furthermore, the post-walnut levels of these three metabolites trended higher in the high urolithin A producers compared to the low urolithin A producers. These mixed results warrant further investigation with a larger study cohort, especially considering that these effects were independent of participant obesity status. While participants with different systemic urolithin A levels may have responded differently to walnut supplementation, our findings do not conclusively establish a definitive role for urolithin A in modifying the fecal levels of putrefaction end-products.

The present study has several limitations. The total participant pool from our clinical trial was unevenly split among BMI groups (n = 12 obese *vs*. n = 27 non-obese). For SCFA analysis, the sample size (n = 21) was relatively small and not equally balanced between BMI subgroups (n = 8 obese *vs*. n = 13 non-obese). Furthermore, our subgroup analyses of SCFAs were limited to 21/39 total participants. When comparing the effects of walnut intake on SCFA levels, alpha diversity, and F/B ratio, it is important to recognize that the former two endpoints were calculated using data from participants that were not included in our SCFA analyses. While the results of our study support the bidirectional association between obesity, dysbiosis, and colonic protein fermentation, it remains unclear whether a causal connection exists between these underlying factors. In addition, our study population had limited generalizability in terms of race, with White individuals making up 64.1% of the sample (n = 25). Of the remaining participants, 20.5% were Asian (n = 8), 12.8% were Black (n = 5), and were 2.6% Hawaiian/Pacific Islander (n = 1).

## Conclusion

We have shown that dietary supplementation with walnut as a primary source of ellagitannins may promote a favorable overall luminal metabolome that is characterized by reduced formation of putrefaction end-products. These effects were especially pronounced in obese individuals with relatively low fecal alpha diversity and higher trending F/B ratio compared to their non-obese counterparts. Together, these data suggest that walnut intake may help to mitigate obesity-related gut dysbiosis. While further research is required to elucidate the role of putrefaction end-products in metabolic and chronic diseases, walnut consumption should be considered for its ability to promote gut health and reduce potential markers of inflammation and dysbiosis in an otherwise healthy clinical cohort.

## Data Availability

Publicly available in a repository:
Statistical analyses, metabolomics data, and sample processing protocols were deposited into the MetaboLights database under accession code REQ20260805222409.

---

**Supplemental Figure 1. The combined effect of urolithin A-producer status and BMI on changes in fecal p-cresol levels during walnut supplementation**. The fecal levels of p-cresol in (A) non-obese and (B) obese individuals, before and after walnut supplementation, were compared to determine whether endogenous urolithin formation correlates with changes in metabolite levels in these groups. Linear mixed models were used to assess the combined effect of BMI and urolithin A producer status (high *vs.* low). Data were log-10 transformed to adjust for numerical skew. A *p*-value less than 0.05 was considered statistically significant (*). Raw values were plotted for ease of visualization, while log-transformed values were used for statistical analysis. Sample sizes for non-obese low and non-obese high-urolithin A were n = 6 and n = 7, respectively. Sample sizes for obese low and obese high-urolithin A were n = 4 and n = 4, respectively.

**Supplemental Figure 2. The differential effects of walnut supplementation on gut microbe abundancy in obese and non-obese volunteers.** Fecal levels of specific microbial genera were determined in non-obese and obese subjects, before and after walnut supplementation. A linear mixed model was used to assess the combined effect of walnut intake and BMI. Changes in the fecal levels of (A) *Peptostreptococcus,* (B) *Clostridium Senso Stricto* 1 and (C) 11 saccharolytic genera [13] are shown, before and after walnut supplementation. Data were log-10 transformed to adjust for numerical skew. A *p*-value less than 0.05 was considered statistically significant (*). Raw values were plotted for ease of visualization, while log-transformed values were used for statistical analysis. Sample sizes were n = 8 and n = 13 for obese and non-obese groups, respectively.

