## Supplemental Tables and Figures for "Effects of Walnut Supplementation on Short-Chain Fatty Acid Levels in Healthy Volunteers"

### Slide 1
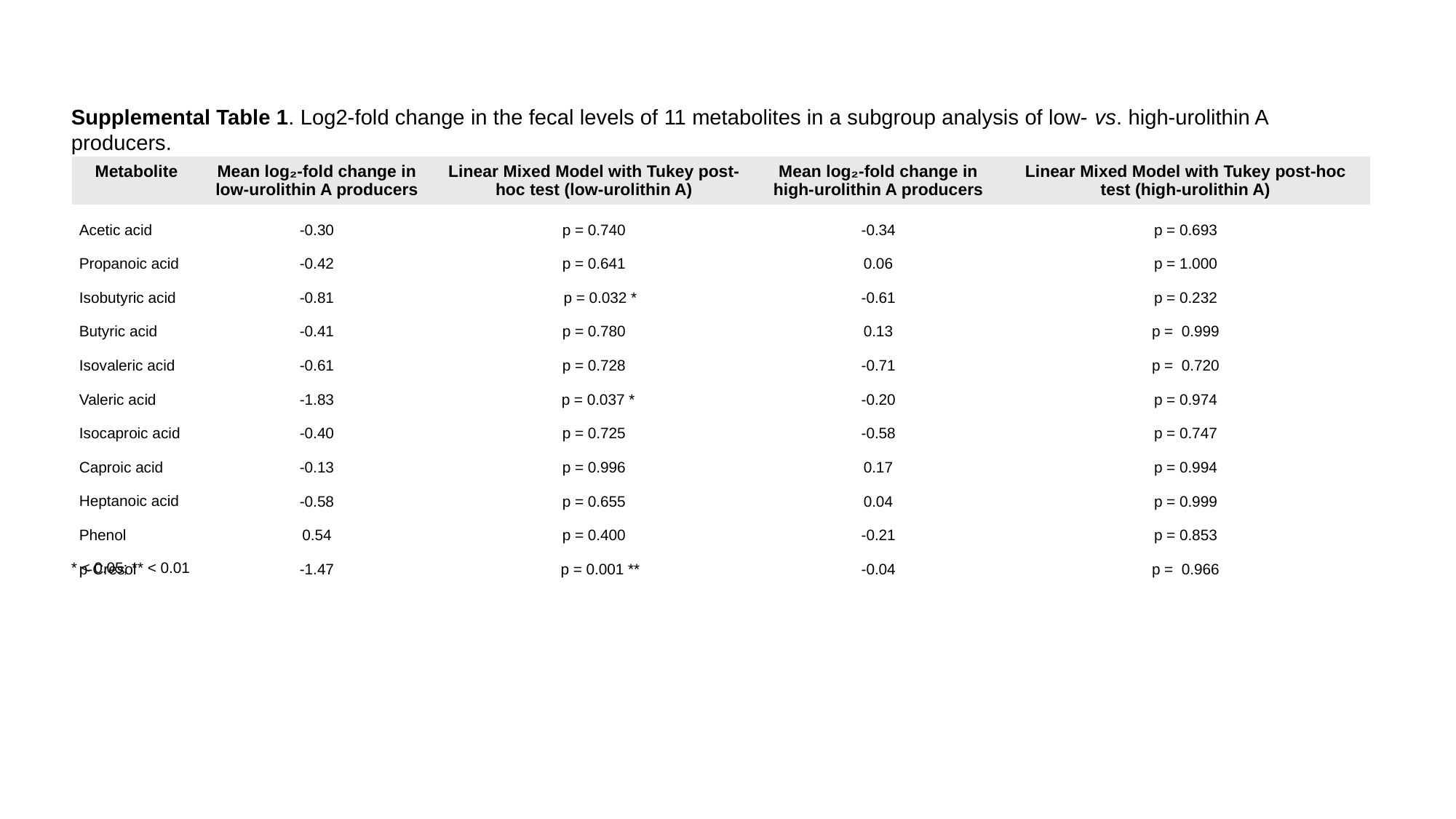

Supplemental Table 1. Log2-fold change in the fecal levels of 11 metabolites in a subgroup analysis of low- vs. high-urolithin A producers.
| Metabolite | Mean log₂-fold change in low-urolithin A producers | Linear Mixed Model with Tukey post-hoc test (low-urolithin A) | Mean log₂-fold change in high-urolithin A producers | Linear Mixed Model with Tukey post-hoc test (high-urolithin A) |
| --- | --- | --- | --- | --- |
| Acetic acid | -0.30 | p = 0.740 | -0.34 | p = 0.693 |
| Propanoic acid | -0.42 | p = 0.641 | 0.06 | p = 1.000 |
| Isobutyric acid | -0.81 | p = 0.032 \* | -0.61 | p = 0.232 |
| Butyric acid | -0.41 | p = 0.780 | 0.13 | p = 0.999 |
| Isovaleric acid | -0.61 | p = 0.728 | -0.71 | p = 0.720 |
| Valeric acid | -1.83 | p = 0.037 \* | -0.20 | p = 0.974 |
| Isocaproic acid | -0.40 | p = 0.725 | -0.58 | p = 0.747 |
| Caproic acid | -0.13 | p = 0.996 | 0.17 | p = 0.994 |
| Heptanoic acid | -0.58 | p = 0.655 | 0.04 | p = 0.999 |
| Phenol | 0.54 | p = 0.400 | -0.21 | p = 0.853 |
| p-Cresol | -1.47 | p = 0.001 \*\* | -0.04 | p = 0.966 |
* < 0.05; ** < 0.01

### Slide 2
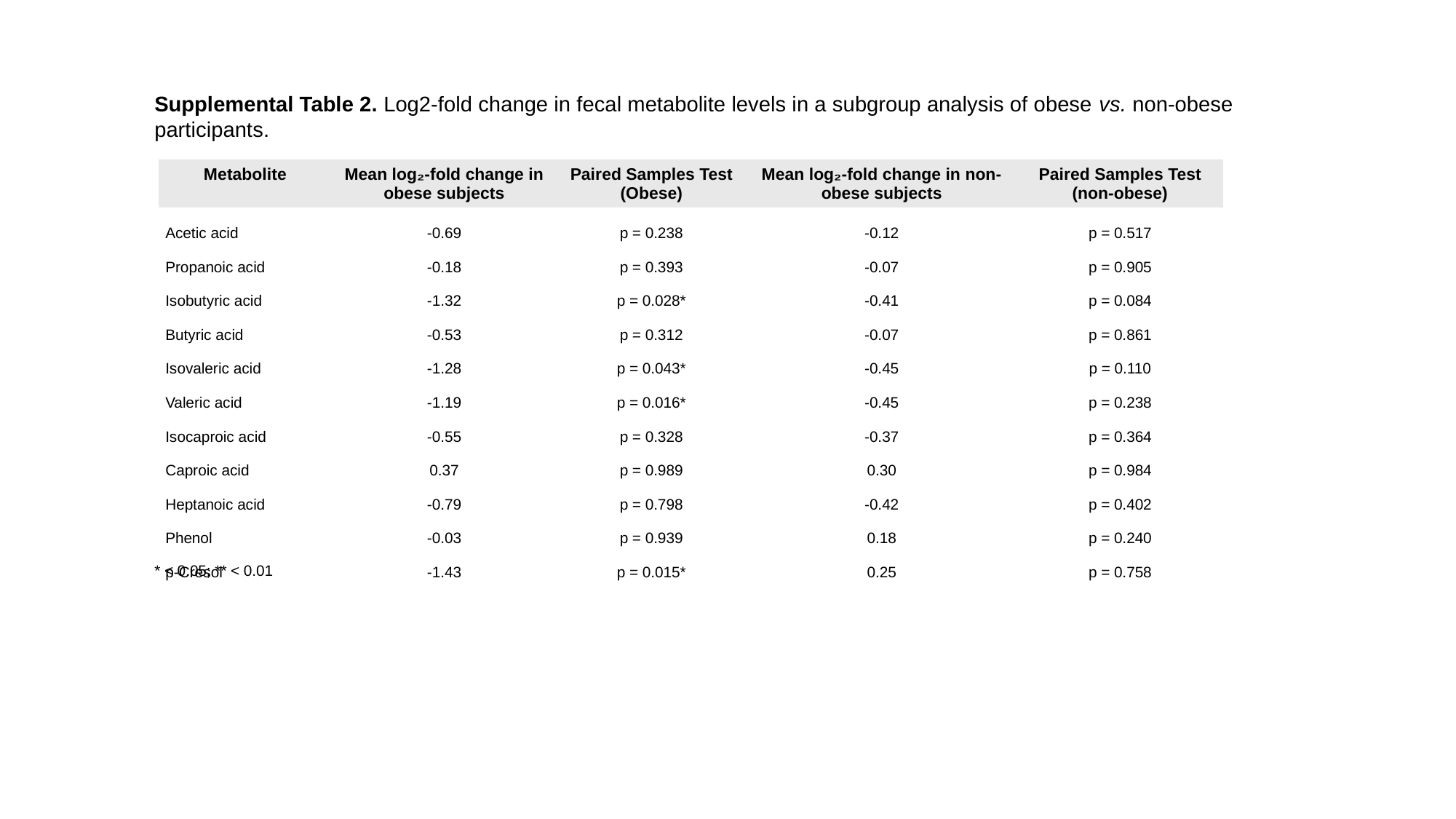

Supplemental Table 2. Log2-fold change in fecal metabolite levels in a subgroup analysis of obese vs. non-obese participants.
| Metabolite | Mean log₂-fold change in obese subjects | Paired Samples Test (Obese) | Mean log₂-fold change in non-obese subjects | Paired Samples Test (non-obese) |
| --- | --- | --- | --- | --- |
| Acetic acid | -0.69 | p = 0.238 | -0.12 | p = 0.517 |
| Propanoic acid | -0.18 | p = 0.393 | -0.07 | p = 0.905 |
| Isobutyric acid | -1.32 | p = 0.028\* | -0.41 | p = 0.084 |
| Butyric acid | -0.53 | p = 0.312 | -0.07 | p = 0.861 |
| Isovaleric acid | -1.28 | p = 0.043\* | -0.45 | p = 0.110 |
| Valeric acid | -1.19 | p = 0.016\* | -0.45 | p = 0.238 |
| Isocaproic acid | -0.55 | p = 0.328 | -0.37 | p = 0.364 |
| Caproic acid | 0.37 | p = 0.989 | 0.30 | p = 0.984 |
| Heptanoic acid | -0.79 | p = 0.798 | -0.42 | p = 0.402 |
| Phenol | -0.03 | p = 0.939 | 0.18 | p = 0.240 |
| p-Cresol | -1.43 | p = 0.015\* | 0.25 | p = 0.758 |
* < 0.05; ** < 0.01

### Slide 3
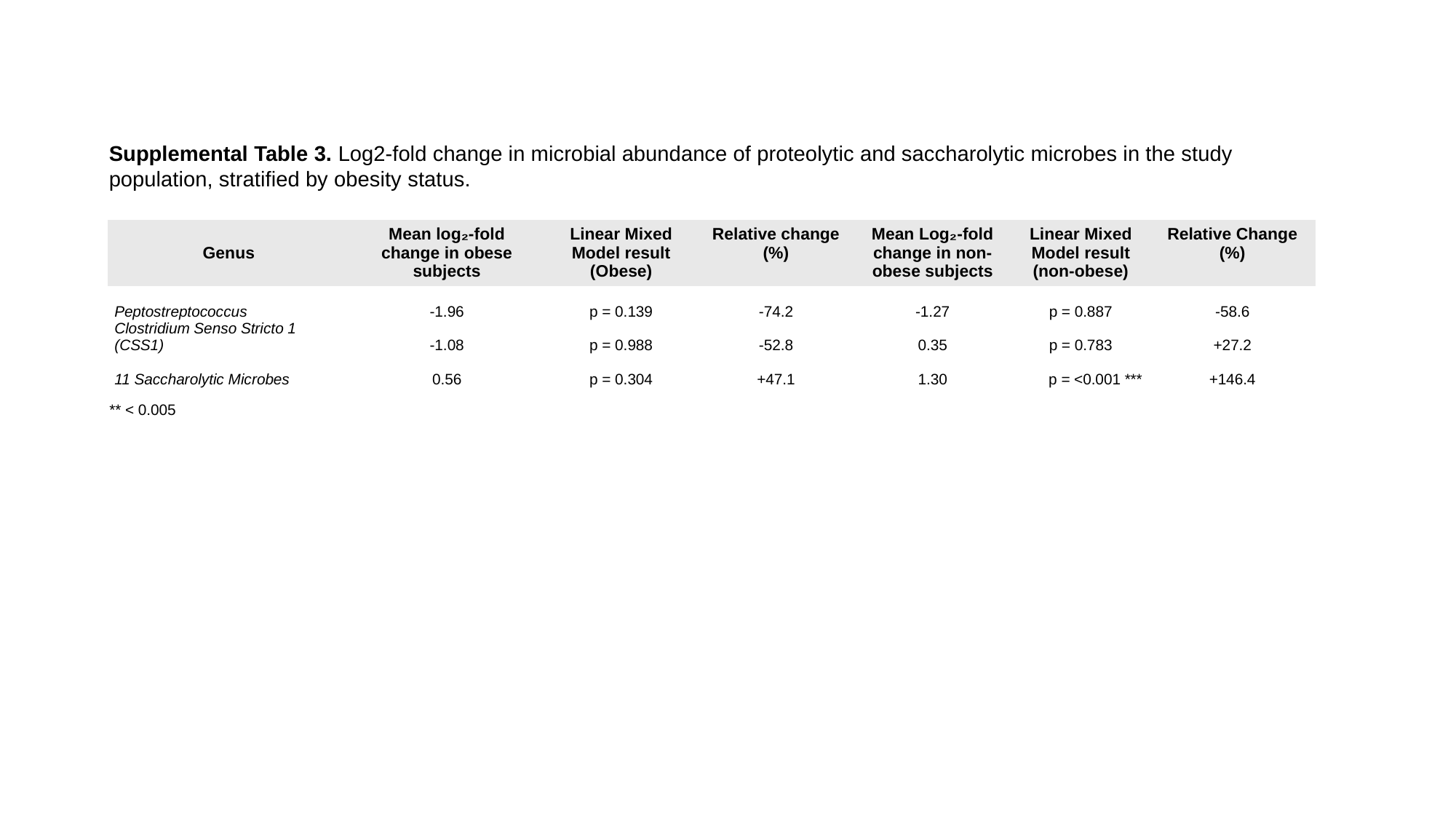

Supplemental Table 3. Log2-fold change in microbial abundance of proteolytic and saccharolytic microbes in the study population, stratified by obesity status.
| Genus | Mean log₂-fold change in obese subjects | Linear Mixed Model result (Obese) | Relative change (%) | Mean Log₂-fold change in non-obese subjects | Linear Mixed Model result (non-obese) | Relative Change (%) |
| --- | --- | --- | --- | --- | --- | --- |
| Peptostreptococcus | -1.96 | p = 0.139 | -74.2 | -1.27 | p = 0.887 | -58.6 |
| Clostridium Senso Stricto 1 (CSS1) | -1.08 | p = 0.988 | -52.8 | 0.35 | p = 0.783 | +27.2 |
| 11 Saccharolytic Microbes | 0.56 | p = 0.304 | +47.1 | 1.30 | p = <0.001 \*\*\* | +146.4 |
** < 0.005

### Slide 4
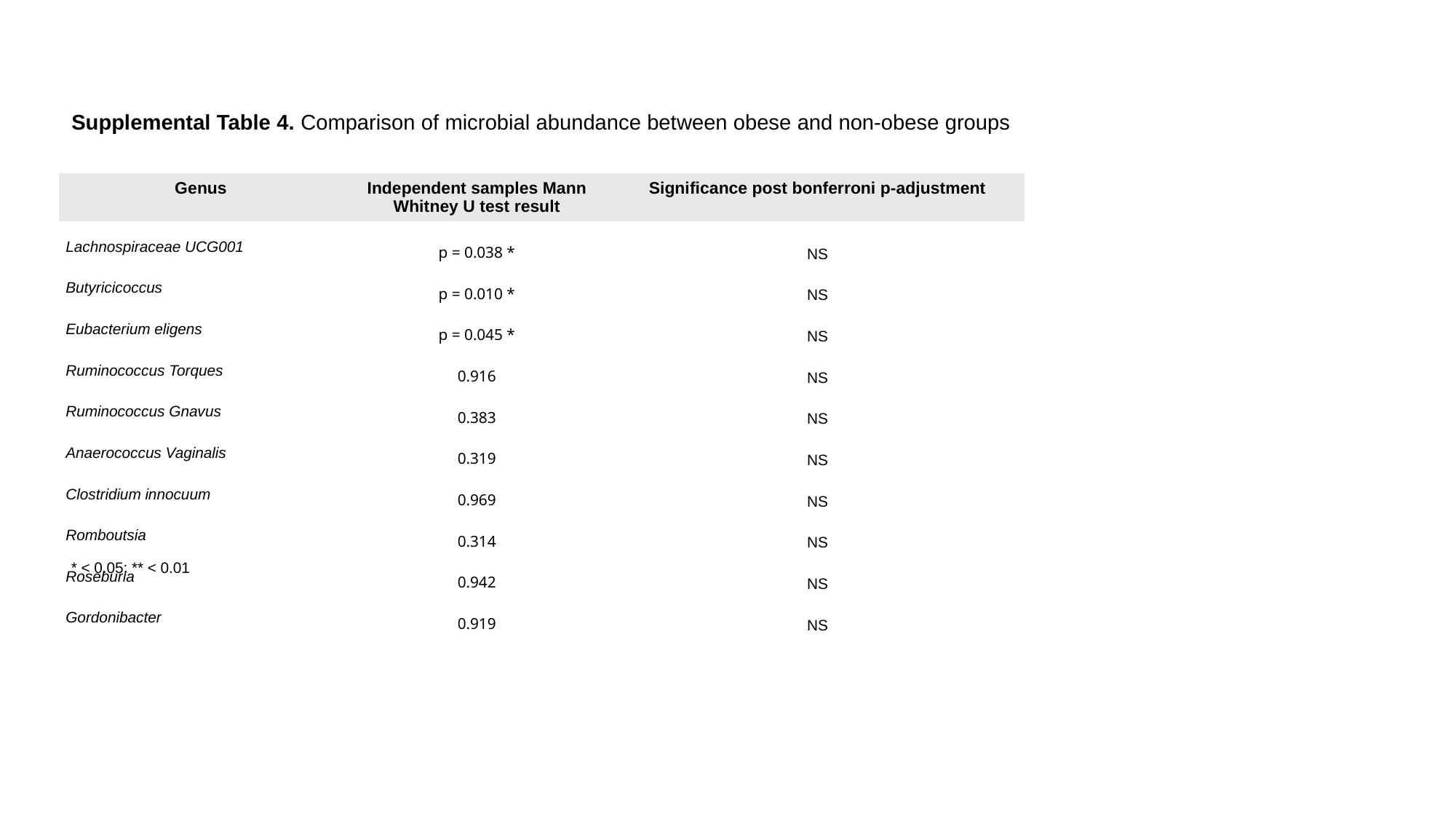

Supplemental Table 4. Comparison of microbial abundance between obese and non-obese groups
| Genus | Independent samples Mann Whitney U test result | Significance post bonferroni p-adjustment |
| --- | --- | --- |
| Lachnospiraceae UCG001 | p = 0.038 \* | NS |
| Butyricicoccus | p = 0.010 \* | NS |
| Eubacterium eligens | p = 0.045 \* | NS |
| Ruminococcus Torques | 0.916 | NS |
| Ruminococcus Gnavus | 0.383 | NS |
| Anaerococcus Vaginalis | 0.319 | NS |
| Clostridium innocuum | 0.969 | NS |
| Romboutsia | 0.314 | NS |
| Roseburia | 0.942 | NS |
| Gordonibacter | 0.919 | NS |
* < 0.05; ** < 0.01

### Slide 5
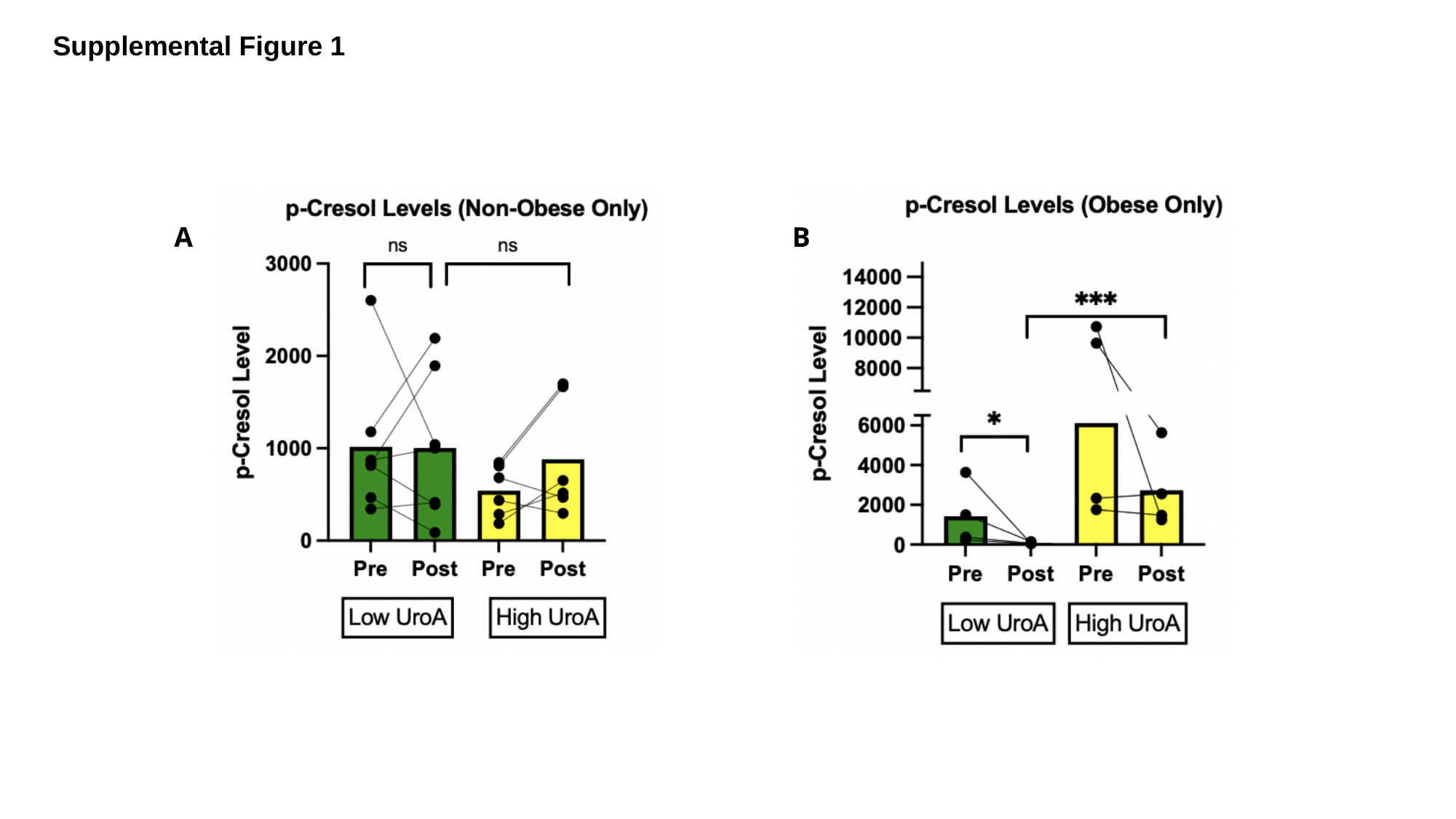

Supplemental Figure 1
B
A

### Slide 6
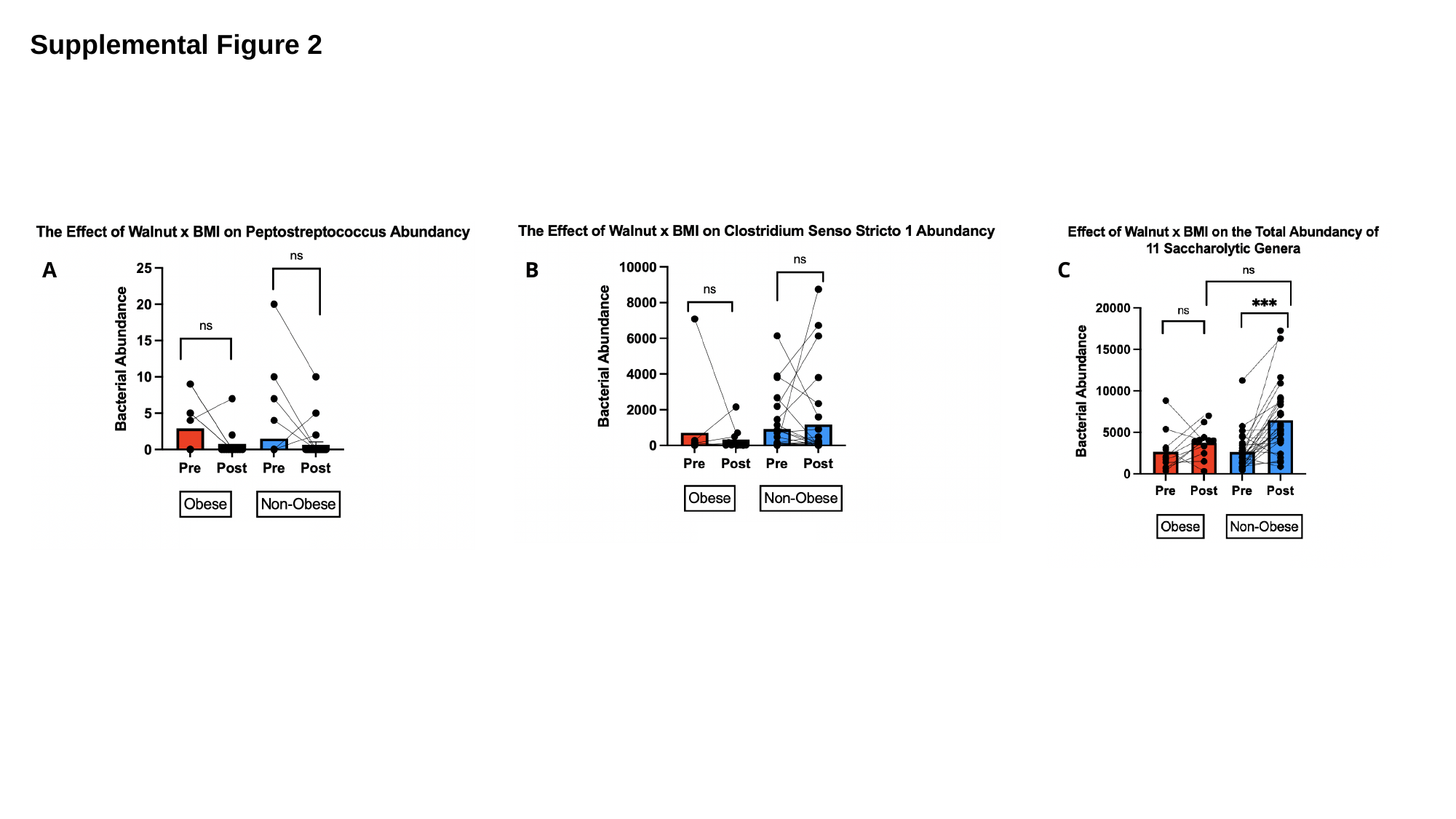

Supplemental Figure 2
A
B
C
